# Severe infections, domain-specific cognitive vulnerability, and future infection risk in older adults

**DOI:** 10.64898/2026.02.17.26346454

**Authors:** Yaqing Gao, Mika Kivimäki, Philipp Frank, Shaun Scholes, Paola Zaninotto, Andrew Steptoe

**Author notes:** Correspondence to: Yaqing Gao, Department of Epidemiology and Public Health, Institute of Epidemiology and Health Care, University College London, 1-19 Torrington Place, London WC1E 7HB, UK.

## Abstract

**Objective:** Severe infections have been implicated in dementia risk, but their associations with detailed patterns of cognitive performance, and whether poorer cognition in turn increases risk for certain infections, remain unclear. We examined bidirectional associations between hospital-treated infections and domain-specific cognitive function in a cohort of older adults.

**Methods:** We analysed data from the English Longitudinal Study of Ageing Harmonised Cognitive Assessment Protocol (ELSA-HCAP), conducted in 2018 and linked to national inpatient records. Pre-HCAP hospital-treated infections were identified from 1997 to 2018; post-HCAP incident infections were ascertained from 2018 to 2024. Cognitive performance was assessed at HCAP using 21 standardised neuropsychological tests summarised into general and four domain-specific scores (executive function, memory, language, and visuospatial ability). Linear regression assessed associations between pre-HCAP hospital-treated infections and standardised cognitive scores; Cox models estimated associations between cognition and risk of incident hospital-treated infections after HCAP. All models were adjusted for sociodemographic, lifestyle, and health covariates.

**Results:** Of 1,159 participants aged ≥65 at HCAP (631 [54.1%] female; mean [SD] age, 75.6 [7.2] years), 351 (30.3%) had a hospital-treated infection before HCAP. Prior hospitalisation for any infection was associated with lower general cognition (β = –0.11 SD, 95% CI –0.21 to –0.02) and poorer executive function (β = –0.19, –0.28 to –0.09), with similar patterns across infection types. Lower respiratory tract infections were additionally associated with poorer memory (β = –0.20, –0.36 to –0.04). Cognitive scores were progressively lower among individuals with more frequent or prolonged infection-related hospitalisations, sepsis, or cardiovascular disease. Prospectively, over a mean (SD) 4.8 (1.9) years of follow-up, 271 incident hospital-treated infections occurred. Each 1-SD higher general cognition was associated with a 36% lower risk of any subsequent hospital-treated infection (HR 0.64, 0.53 to 0.78), and with consistent associations across cognitive domains for all-cause and bacterial infections. Executive function alone showed a strong association with viral infections, especially COVID-19 (HR 0.59, 0.44 to 0.80).

**Conclusion:** Severe infections were primarily associated with poorer executive function. Conversely, cognitive vulnerability across multiple domains was associated with increased susceptibility to infections requiring hospital care, while poorer executive function was specifically associated with viral infection risk. These findings support a reinforcing infection–cognition cycle in later life and cognitively tailored infection-prevention strategies.

## 1.1 Introduction

Severe infection has increasingly been implicated as a potential contributor to dementia development. Recent large-scale registry studies have reported consistent associations between a wide range of hospital-treated infections—including bacterial, viral, central nervous system (CNS), and extra-CNS infections such as pneumonia and urinary tract infections—and increased dementia risk.^1–3^ However, studies relying on dementia diagnoses from health registries are prone to ascertainment bias, as individuals with severe infections are more likely to engage with healthcare services and, therefore, more likely to have dementia detected. Moreover, health registries may fail to capture milder cases. Assessing cognitive performance in population-based cohorts provides an intermediate outcome less influenced by healthcare utilisation and may capture early cognitive changes preceding dementia diagnosis.

Few studies have examined hospital-treated infections in relation to cognitive performance. Most have focused on single infections, typically pneumonia or COVID-19, and consistently report poorer global cognitive function following infection.^4–7^ Whether these associations extend to a broader range of infections remains unclear. Reliance on global cognitive scores also limits detection of subtle or heterogeneous deficits. Registry-based studies have reported stronger associations between hospital-treated infections and vascular dementia than with Alzheimer’s disease,^1,2^ suggesting that infections may preferentially affect cerebrovascular pathways. Domain-specific assessments are therefore needed to shed light on potential underlying mechanisms. Only one study has examined multiple common infections—including lower respiratory, urinary, and skin or soft tissue infections—treated in both primary and secondary care in relation to cognitive decline across domains such as reaction time, memory, and fluid intelligence, and it reported no robust associations.^8^ However, this study is limited by the relatively young sample (mean age 56 years), the high prevalence of milder infections not requiring hospital treatment, and the use of brief, computerised, non-standardised cognitive assessments that may have introduced measurement bias.

This association may act bidirectionally, whereby cognitive impairment increases vulnerability to serious infections, creating a potential vicious cycle. Stronger infection-dementia associations over shorter follow-up periods may partly reflect infections occurring during the preclinical stage of dementia.^1,2^ Few studies have directly examined this reverse association; most evidence comes from COVID-19 research showing that individuals with dementia or cognitive impairment have a higher risk of infection-related hospitalisation,^9 10,11^ likely due to compromised self-care. Whether such vulnerability extends to non-COVID-19 infections, and which cognitive domains primarily contribute to this risk, remains uncertain.

In this study, we used detailed multidomain neuropsychological assessments to examine whether a history of hospital-treated infection was associated with poorer general and domain-specific cognition, whether cognitive performance predicted subsequent incident infection risk, and whether these associations varied by pathogen type, infection site, severity, or participant characteristics.

## 1.2 Material and methods

### 1.2.1 Study population

The English Longitudinal Study of Ageing (ELSA) is an ongoing, nationally representative cohort of adults aged ≥50 years living in England.^12^ Established in 2002, ELSA follows participants biennially through computer-assisted interviews and self-completion questionnaires collecting detailed information on social, economic, health, and psychological factors. Periodic refreshment samples are added to maintain representativeness of the English household population aged ≥50 years.

The Harmonised Cognitive Assessment Protocol (HCAP) is a substudy conducted in 2018 to provide detailed cognitive assessments among older ELSA participants.^13^ Eligible participants were core ELSA members aged ≥65 years who had completed at least one self-interview in 2014–2015 (wave 7) or 2016–2017 (wave 8). HCAP was administered across the full range of cognitive abilities, with oversampling of participants with lower cognitive scores identified in the most recent ELSA waves. Face-to-face HCAP interviews were conducted between 15 January and 8 April 2018; because individual interview dates were unavailable due to disclosure restrictions, a midpoint date (26 February 2018) was assigned as the interview date for all participants.

Of 1,684 eligible participants, 1,273 completed the HCAP interview (response rate 75.6%), and 1,173 consented to and were successfully linked with Hospital Episode Statistics (HES) records, which capture all admissions to publicly funded hospitals in England. For cross-sectional analyses of pre-HCAP hospital-treated infection and HCAP cognitive function, participants with missing covariate data (n=14) were excluded, yielding a final sample of 1,159. For prospective analyses, participants with any pre-HCAP hospital-treated infection (n=351) were further excluded, resulting in 808 participants. A flowchart of participant selection and study design is shown in appendix figure S1.

### 1.2.2 Hospital-treated infection

Hospital-treated infection was defined as having at least one infection diagnosis recorded in the HES-Admitted Patient Care data before HCAP. Diagnoses were identified using International Classification of Diseases, Tenth Revision (ICD-10) codes from a list developed by Sipilä et al,^1^ comprising 925 infection-related codes. ICD-based identification of serious infections in administrative data has previously shown high validity, with positive predictive values typically around 80–90%.^14^

Infections were further classified by common pathogen type (bacterial or viral) and major sites (lower respiratory tract, urinary tract, or gastrointestinal); rarer types were included only in the overall “any infection” category. Infection burden was defined as the number of infection-related hospital admissions (1, 2, or ≥3 episodes). Infection severity was determined by the longest hospital stay across all infection episodes (same-day discharge, 1–3 days, or ≥4 days) and by the presence of sepsis, defined as infection complicated by life-threatening organ dysfunction.^15^

### 1.2.3 Cognitive function

The HCAP battery included a comprehensive set of standardized neuropsychological tests assessing cognitive domains commonly affected by ageing. Tests included the Mini-Mental State Examination (MMSE); Consortium to Establish a Registry for Alzheimer’s Disease (CERAD) Word List Learning, Recall, and Recognition; Logical Memory (*Brave Man* story from the East Boston Memory Test and *Anna Thompson* story from the Wechsler Memory Scale); CERAD Constructional Praxis; Symbol Digit Modalities Test; Number Series; Raven’s Standard Progressive Matrices; and Trail Making Tests A and B, among others (21 tests in total). For all tests, higher scores indicated better cognitive performance, except the Trail Making Tests, which measured total time to complete the task; their scores were inverted so that higher values reflected better performance. To facilitate comparison across cognitive measures, all scores were standardized (mean⍰= ⍰0, SD⍰= ⍰1).

Four domain-specific factor scores (executive function, memory, language, and visuospatial ability) were derived using confirmatory factor analysis (CFA) based on the cognitive test data, with missing values imputed using a block-sequential chained approach incorporating sociodemographic and health covariates from ELSA and HCAP.^16^ A hierarchical CFA was then applied to combine the four domain factors into a higher-order general cognitive factor. All factor scores were standardized (mean⍰= ⍰0, SD⍰= ⍰1). Table S1 summarises the cognitive tests, their assigned domains, and the proportion of missing data.

### 1.2.4 Covariates

Covariates included established risk factors and comorbidities for severe infection and dementia:^17–20^ age (at HCAP participation), sex (male vs female), quintiles of total net household wealth (total assets minus debts), quintiles of the Index of Multiple Deprivation (IMD, a small-area composite measure of socioeconomic disadvantage based on income, employment, education, health, crime, housing, and environment), educational attainment (<O level, O level, ≥A level), living status (alone/co-residing), loneliness (three-item scale; score ≥6 indicating loneliness^21^), smoking status (past or current vs never), and alcohol consumption (≥5 vs <5 days/week). The most recent measures available across ELSA waves prior to HCAP were used to minimize misclassification.

Health conditions included history of diabetes, hypertension, cardiovascular disease (angina, myocardial infarction, heart failure, or stroke), chronic lung disease, and psychiatric conditions, identified from self-reports in HCAP or prior ELSA waves and linked HES records.

Tables S2–S3 describe variable definitions, categorizations, and data sources. Missing covariate data were minimal (1% in total; table S4), and complete case analysis was applied.

### 1.2.5 Statistical analysis

Baseline sociodemographic and health characteristics were summarised for the total sample and stratified by pre-HCAP hospital-treated infection status. Categorical variables were reported as counts and weighted percentages, and continuous variables as weighted means with standard deviations, accounting for differential sampling and participation probabilities in HCAP.

For the cross-sectional analysis, multivariable-adjusted linear regression models were used to examine the associations between pre-HCAP hospital-treated infections (any infection, by pathogen, and by infection site) and cognitive performance measured in HCAP. The primary outcomes were general and domain-specific cognitive factor scores. Secondary analyses examined associations with individual cognitive test scores (n = 21), with multiple comparisons addressed using false discovery rate correction. Dose-response relationships were evaluated for all infections by infection burden and severity in relation to the general and domain-specific cognitive factor scores, with participants without any hospital-treated infection as the reference group.

For the prospective analysis, multivariable-adjusted Cox proportional hazards models were used to examine the association between cognitive function measured at HCAP and subsequent risk of incident hospital-treated infection. Schoenfeld residuals indicated no violation of the proportional hazards assumption (figure S2). The primary exposures were general and domain-specific cognitive factor scores, and the secondary exposures were individual test scores. Participants were followed from their HCAP assessment until the incident infection-related hospital admission, death, or the most recent available linkage date in HES (31 March 2024), whichever occurred first. Follow-up time was used as the underlying timescale. Analyses were also conducted separately by pathogen type and site, and for COVID-19 infections.

For both cross-sectional and prospective analyses, stratified analyses were performed to assess whether associations between all infections and the general cognitive factor score differed by sociodemographic and health subgroups, with p-values for interaction derived from likelihood ratio tests. All models were adjusted for predefined covariates and incorporated HCAP survey weights.^16^

All statistical analyses were performed using R (version 4.4.1).

## 1.3 Results

Among 1,159 HCAP participants aged ≥65 years included in the cross-sectional analysis (631 [54.1%] female; mean [SD] age, 75.6 [7.2] years), 351 (30.3%) had at least one hospital-treated infection before HCAP (table 1). Participants with prior hospital-treated infection were older, more often male, and more likely to have lower educational attainment, lower household wealth, and to live in more socioeconomically deprived areas. They were also more likely to live alone and report loneliness, whereas smoking and alcohol consumption patterns were similar between groups. Participants with prior hospital-treated infections had also a higher prevalence of all examined health conditions, particularly chronic lung disease (30.2% vs 11.8%) and cardiovascular disease (40.1% vs 19.5%).

**Table 1.** Baseline characteristics of participants overall and by infection history before HCAP.

| Characteristic | Overall | Any hospital-treated infection | No hospital-treated infection |
| --- | --- | --- | --- |
| N | 1159 | 351 | 808 |
| Age, mean (SD) | 75.6 (7.2) | 77.0 (7.7) | 75.1 (6.9) |
| Female, n (%) | 631 (54.1) | 187 (49.8) | 444 (55.7) |
| Living alone, n (%) | 369 (29.3) | 130 (35.3) | 239 (27.1) |
| Loneliness, n (%) | 228 (19.5) | 78 (20.7) | 150 (19.0) |
| Education |  |  |  |
| <O-level | 519 (43.8) | 173 (46.5) | 346 (42.8) |
| O-level | 288 (24.5) | 92 (24.8) | 196 (24.4) |
| ≥A-level | 352 (31.8) | 86 (28.7) | 266 (32.9) |
| Wealth quintile <sup>a</sup>, n (%) |  |  |  |
| Quintile 1 (Lowest wealth) | 278 (23.8) | 103 (28.9) | 175 (21.9) |
| Quintile 2 | 278 (21.9) | 88 (21.0) | 190 (22.3) |
| Quintile 3 | 233 (19.9) | 75 (23.1) | 158 (18.7) |
| Quintile 4 | 206 (17.8) | 58 (16.9) | 148 (18.1) |
| Quintile 5 (Highest wealth) | 164 (16.6) | 27 (10.2) | 137 (19.0) |
| IMD quintile <sup>b</sup>, n (%) |  |  |  |
| Quintile 1 (Least deprived) | 245 (20.7) | 56 (14.9) | 189 (22.8) |
| Quintile 2 | 282 (23.1) | 83 (23.3) | 199 (23.1) |
| Quintile 3 | 254 (23.4) | 75 (23.8) | 179 (23.2) |
| Quintile 4 | 217 (18.4) | 82 (24.3) | 135 (16.3) |
| Quintile 5 (Most deprived) | 161 (14.4) | 55 (13.8) | 106 (14.6) |
| Past/current smoking, n (%) | 760 (65.1) | 234 (65.5) | 526 (65.0) |
| Drinking $\geq 5$ days/week, n (%) | 226 (20.3) | 66 (20.7) | 160 (20.1) |
| Diabetes, n (%) | 222 (18.3) | 89 (23.3) | 133 (16.4) |
| Hypertension, n (%) | 753 (63.0) | 271 (73.0) | 482 (59.4) |
| Chronic lung disease, n (%) | 204 (16.7) | 103 (30.2) | 101 (11.8) |
| Cardiovascular disease, n (%) | 346 (24.9) | 162 (40.1) | 184 (19.5) |
| Psychiatric condition, n (%) | 224 (20.0) | 81 (25.6) | 143 (18.0) |

SD, standard deviation; IMD, Index of Multiple Deprivation. All estimated means, standard deviations, and (column) proportions were weighted to account for differential sampling and participation probabilities at HCAP.
<sup>a</sup> Net total wealth measured at the benefit unit level (i.e., a single adult or a couple, plus any dependent children, sharing finances). The sum of savings, investments, physical wealth and housing wealth after financial debt and mortgage debt has been subtracted.
<sup>b</sup> Index of Multiple Deprivation: Area-level measure of socioeconomic deprivation based on participants' postcode, incorporating income, employment, education, health, crime, housing, and environment.

Overall, prior hospital-treated infection was consistently associated with lower general cognition and executive function across minimally and fully adjusted models (table S5). In the fully adjusted analysis, having any hospital-treated infection was associated with a 0.11 SD lower general cognitive factor score compared with no infection (β = –0.11, 95% CI –0.21 to – 0.02; figure 1), while among cognitive domains only executive function remained significantly associated (β = –0.19, 95% CI –0.28 to –0.09). At the test level, this was reflected in poorer performance on the MMSE (global cognition) and three of seven executive function tasks. When examined by pathogen, bacterial infections showed a similar but stronger pattern, for example for executive function (β = –0.24, 95% CI –0.35 to –0.14), with significant associations across five of seven executive tests, whereas viral infections showed little evidence of association with any cognitive domain. Both general and executive domains showed consistent associations across infection sites. Lower respiratory tract infections (LRTI) were additionally associated with lower memory factor scores (β = –0.20, 95% CI –0.36 to –0.04), particularly in recognition memory tests.

**Figure 1.**
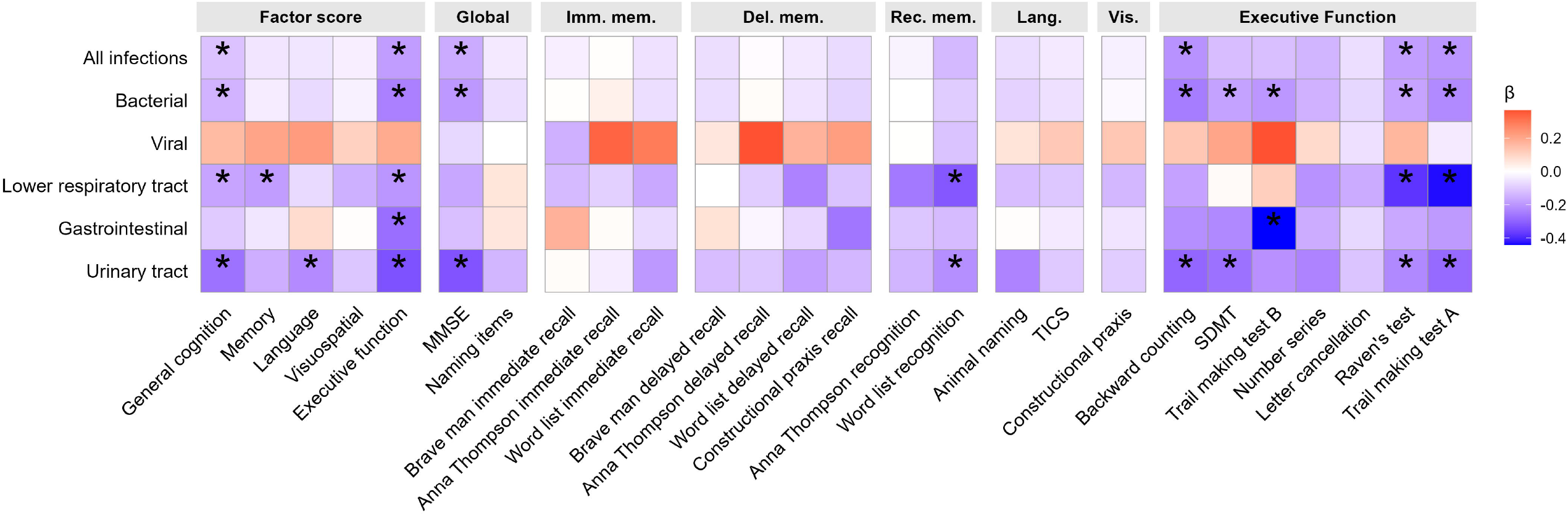
Associations between pre-HCAP hospital-treated infection and HCAP cognitive function. Imm.mem., Immediate memory; Del.mem., Delayed memory; Rec.mem., Recognition memory; Lang., Language; Vis., Visuospatial; MMSE, Mini-Mental State Examination; TICS, Telephone Interview for Cognitive Status; SDMT, Symbol Digit Modalities Test. Associations of pre-HCAP hospital-treated infection with factor scores and individual cognitive test scores by domain. Models were estimated using linear regression, adjusted for demographics, socioeconomic, lifestyle, and health covariates and weighted to account for differential sampling and participation probabilities at HCAP. For factor scores, * denotes p < 0.05; for individual tests, * denotes FDR-corrected p < 0.05.

Compared with participants without pre-HCAP infection-related admissions, those with ≥3 admissions showed significantly lower general cognitive factor scores (β = –0.27, 95% CI –0.46 to –0.08; figure 2), while the associations for 1 or 2 admissions were weaker and not statistically significant. Similarly, a significant association was observed only for hospitalisations with the longest stay ≥4 days (β = –0.16, 95% CI –0.29 to –0.04), and participants with sepsis had the lowest general cognitive scores (β = –0.32, 95% CI –0.60 to –0.04). Across cognitive domains, most associations remained non-significant except for executive function, which demonstrated a clearer dose-response relationship.

**Figure 2.**
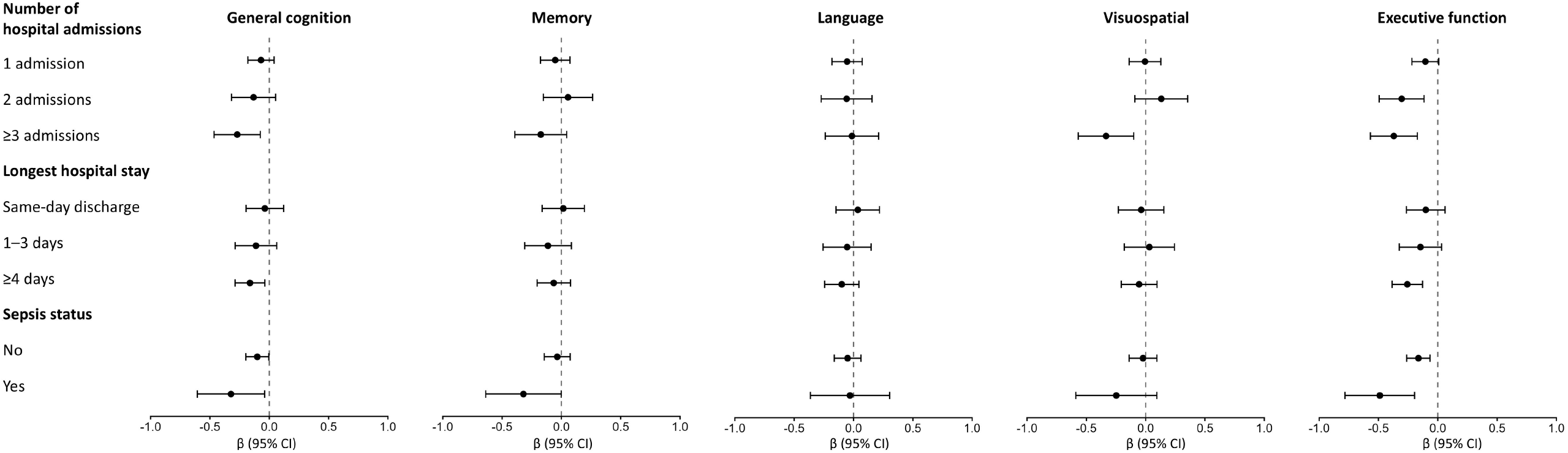
Associations between pre-HCAP infection burden and severity and HCAP cognitive function. CI, confidence intervals. Associations between pre-HCAP hospital-treated infection characteristics and general and domain-specific cognitive factor scores at HCAP. Infection burden was defined as the number of infection-related hospital admissions, infection severity by the longest infection-related hospital stay and whether any infection admission involved sepsis. All estimates are compared with participants with no hospital-treated infections. Models were estimated using linear regression, adjusted for demographic, socioeconomic, lifestyle, and health covariates, and weighted to account for differential sampling and participation probabilities at HCAP.

In analyses examining the opposite temporal direction, 808 participants without prior hospital-treated infection (449 [55.2%] female; mean [SD] age, 76.3 [7.0] years) were followed for up to 6 years after HCAP, during which 271 had an incident hospital-treated infection. Associations were consistent across minimally and fully adjusted models (table S6). In the fully adjusted analysis, each SD higher general cognitive factor score was associated with a 36% lower risk of developing any hospital-treated infection after HCAP (HR 0.64, 95% CI 0.53 to 0.78; figure 3). Similar protective associations were observed across multiple cognitive domains, including memory (HR 0.71, 95% CI 0.60 to 0.86), language (HR 0.77, 95% CI 0.65 to 0.93), and most strongly, executive function (HR 0.67, 95% CI 0.56 to 0.82), with consistent associations across 3 of 7 executive tests. Patterns were similar for bacterial and urinary tract infections. For viral infections, associations were limited to general and executive function, the latter showing the strongest association with COVID-19: each 1-SD higher executive score was linked to a 41% lower risk of COVID-19 hospitalisation (HR 0.59, 95% CI 0.44 to 0.80), consistent across 5 of 7 executive tests.

**Figure 3.**
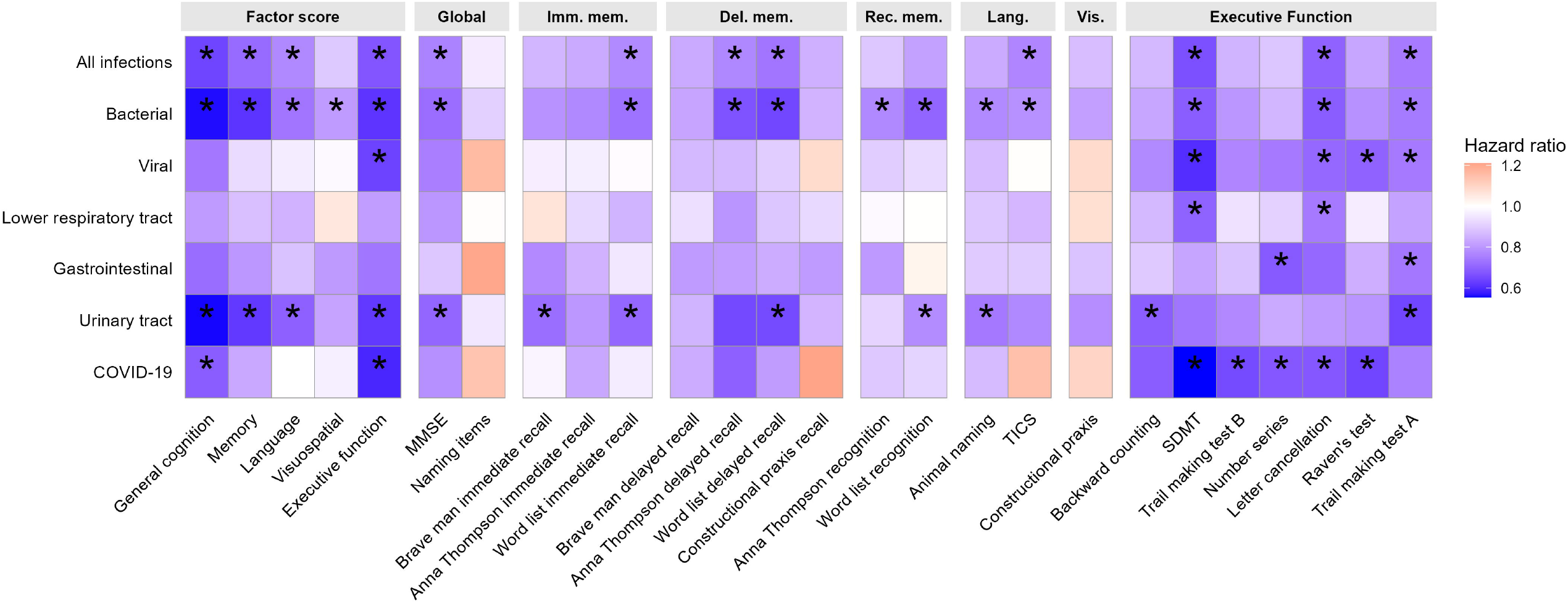
Associations between cognitive function in HCAP and risk of subsequent hospital-treated infection. Imm.mem., Immediate memory; Del.mem., Delayed memory; Rec.mem., Recognition memory; Lang., Language; Vis., Visuospatial; MMSE, Mini-Mental State Examination; TICS, Telephone Interview for Cognitive Status; SDMT, Symbol Digit Modalities Test. Associations of HCAP cognitive factor scores and individual cognitive test scores with incident hospital-treated infection. Hazard ratios were estimated using Cox regression models adjusted for demographic, socioeconomic, lifestyle, and health covariates, and weighted to account for differential sampling and participation probabilities at HCAP. For factor scores, * denotes p < 0.05; for individual tests, * denotes FDR-corrected p < 0.05.

Associations between pre-HCAP hospital-treated infection and HCAP general cognitive function were generally consistent across subgroups (p_interaction_ >0.05; figure 4). Stronger associations were observed among participants in lower household wealth quintiles, those who drank alcohol less frequently (<5 days/week; β = –0.15, 95% CI –0.26 to –0.04 vs β = 0.09, 95% CI – 0.12 to 0.30 for ≥5 days/week), and those with a history of cardiovascular disease (β = –0.24, 95% CI –0.40 to –0.08 vs β = –0.03, 95% CI –0.15 to 0.08). Prospectively, higher general cognitive function was consistently associated with a lower risk of subsequent hospital-treated infection across subgroups (figure S3). As no correction for multiple comparisons was applied, interaction analyses should be interpreted as hypothesis-generating.

**Figure 4.**
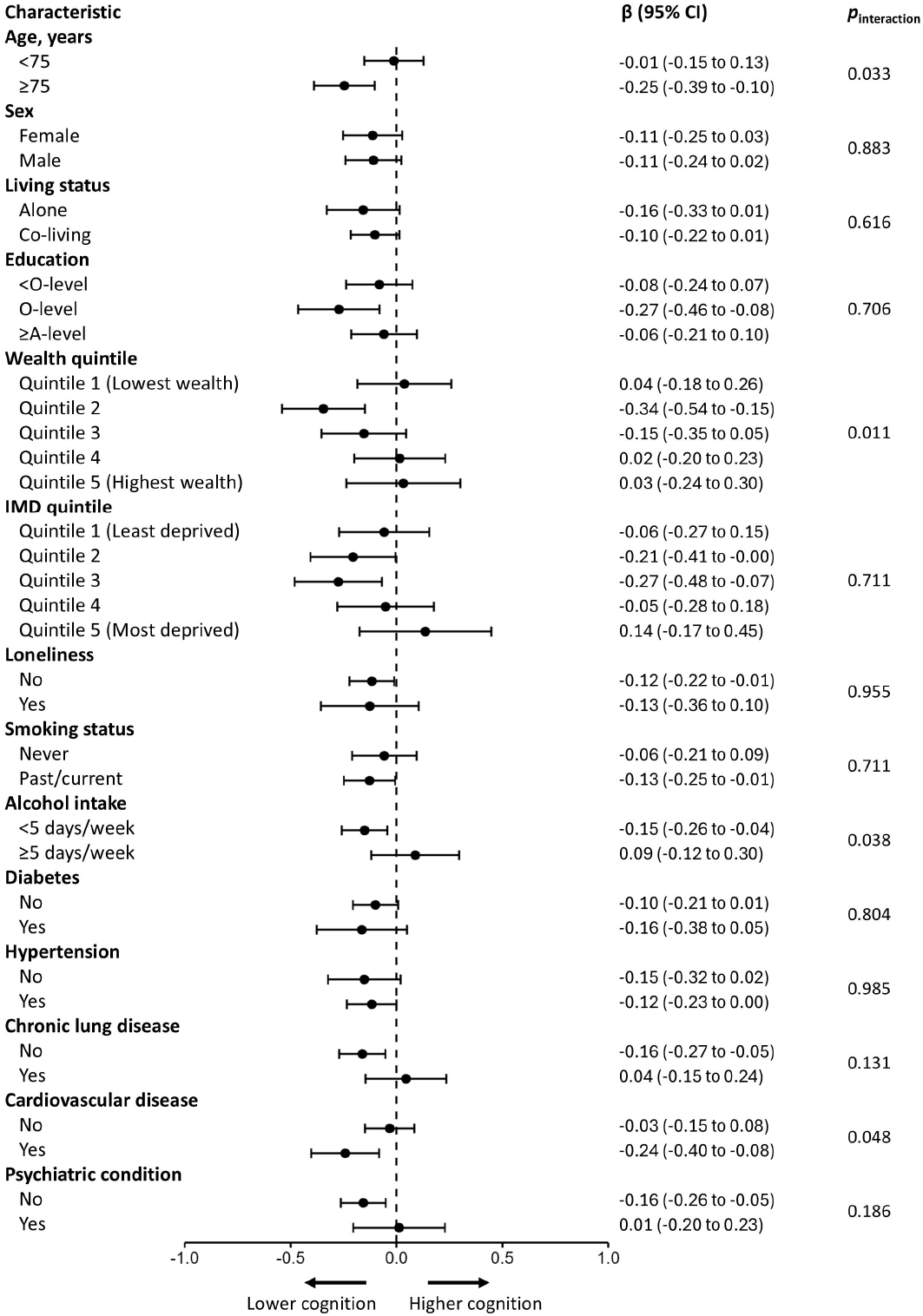
Association between pre-HCAP hospital-treated infection and HCAP general cognitive function by subgroup. CI, confidence intervals; IMD, Index of Multiple Deprivation. Associations of any hospital-treated infection before HCAP with general cognitive factor scores, stratified by demographic, socioeconomic, lifestyle, and health characteristics. Models were adjusted for all listed covariates, and weighted to account for differential sampling and participation probabilities. p_interaction_ values were obtained using likelihood ratio tests.

## 1.4 Discussion

We observed bidirectional associations between hospital-treated infections and cognition, with distinct domain–infection patterns in each direction. Prior infections were linked mainly to poorer executive function, with LRTI also being associated with poorer memory. Associations were stronger among individuals with greater infection burden, higher severity, cardiovascular disease, and lower socioeconomic status. In contrast, lower baseline cognitive performance across multiple domains—including executive, memory, and language—was associated with higher risk of subsequent bacterial infections, whereas only executive function was associated with future viral infections, including COVID-19.

Our findings suggest that systemic inflammation, rather than specific pathogens, underlies the link between infections and cognitive impairment. Two lines of evidence support this inference. First, we observed similar associations across multiple infection types, aligning with evidence that diverse infections are linked to higher dementia risk.^1–3^ This pattern appears consistent across the life course: a Danish nationwide study of over 160,000 young men (mean age 19) found that a wide range of infections were associated with lower cognitive ability.^22^ Although we observed no association between viral infections and cognition, this likely reflects limited power (only 19 viral hospitalisations before HCAP). Prior studies have shown that hospital-treated viral infections are associated with higher dementia risk, varicella–zoster vaccination with lower dementia risk,^23,24^ and seropositivity for herpes simplex virus 2 or cytomegalovirus with poorer executive function.^25^ Second, we identified a clear dose-response pattern: greater infection burden and more severe infections were associated with progressively lower cognitive performance. This pattern aligns with previous findings that repeated infections, sepsis, and hospital-treated (but not primary care-treated) infections are particularly linked to higher dementia risk and faster cognitive decline.^1,2,7^

We present the first evidence that prior hospital-treated infections show a domain-specific cognitive profile, with the strongest and most consistent associations observed for executive function, while other domains showed little evidence of association. The exception was LRTI, which was additionally associated with poorer memory, and this appeared unidirectional, as baseline memory did not predict future LRTI risk. This pattern is consistent with reports that hospital-treated COVID-19 is linked to memory and executive function but not language.^7^ Domain-specific associations across different infections have also been suggested in proteomic studies: proteins linked to prior human herpesvirus infection relate most strongly to executive domains, whereas influenza-related proteins show clearer associations with memory decline.^26^ The additional memory association seen for LRTI may reflect its pronounced propensity to reduce oxygen supply. Inflammation and fluid accumulation within the alveoli impair gas exchange and limit oxygen diffusion into the bloodstream, and memory-critical brain regions such as the hippocampus and entorhinal cortex are particularly sensitive to reduced oxygen availability.^27,28^

The stronger associations with executive function point to a vascular pathway linking severe infection to cognitive impairment. Specific mechanistic explanations for the link between infection and vascular-mediated cognitive decline include endothelial dysfunction and thrombosis secondary to acute inflammation, infection-related platelet aggregation, and accelerated atherogenesis.^29^ Systemic inflammation can destabilise atherosclerotic plaques and increase the risk of clot formation,^30^ consistent with evidence that hospital-treated infections increase the risk of myocardial infarction and stroke.^31,32^ It may also directly disrupt the blood-brain barrier and impair microcirculation.^33^ Executive function depends on frontostriatal circuits, which contain long, metabolically demanding white-matter tracts supplied by small penetrating arteries with limited autoregulatory capacity,^34^ making them particularly vulnerable to ischaemic stress. Accordingly, executive function impairment is a recognised hallmark of cerebral small-vessel disease and vascular cognitive impairment,^35^ and neuropsychological comparisons consistently show greater executive function deficits in vascular dementia than in Alzheimer’s disease.^36,37^ This also aligns with our observation that infection-cognition associations were stronger in individuals with cardiovascular disease. Prior studies similarly show that infection-related dementia risk^38^ or slowed processing speed^39^ is greatest among those with poorer cardiovascular health.

Cognitive domains showed distinct associations with subsequent infection risk: poorer performance across all domains predicted higher all-cause and bacterial infection risk, whereas only executive function was associated with future viral infection, largely reflecting COVID-19 (86 of 119 incident viral infections). The lack of associations between pre-HCAP infections and memory or language, together with null findings for pre-HCAP viral infections across all domains, supports the direction that lower cognition precedes higher infection risk. Evidence for this direction has been limited, apart from nationwide data showing that dementia increases the risk of hospital-treated respiratory, urinary and gastrointestinal infections,^40^ and that slower reaction time predicts higher mortality from lower respiratory tract infections, including COVID-19.^41,42^ The domain specificity we observed may reflect the behavioural demands of preventing different infections. Avoiding COVID-19 and other viral exposures requires rapidly adaptive behaviours, such as responding to changing public-health guidance, that rely particularly on executive function skills. In contrast, prevention of other infections relies more on routine self-care, including hygiene, medication adherence and timely symptom reporting, which involve a broader set of cognitive abilities, including memory and language.

Using nationwide inpatient data linked for the study cohort and standardized, multidomain cognitive assessments, this study provides a comprehensive evaluation of infection-cognition associations. The combined use of pre-HCAP infection history and prospective follow-up for subsequent infections also provides insight into the directionality of some associations. Several limitations should be acknowledged. Our data capture only inpatient-treated infections, with milder cases managed in primary care or at home unavailable, limiting our ability to distinguish incidence from severity. However, previous studies do not support an association between milder infections and dementia.^2,43^ Statistical power was limited for less common infection subtypes, particularly viral infections. Among invited participants, those who completed HCAP tended to have higher education, better cognition and fewer functional impairments;^44^ although survey weights were applied to partially address this participation bias, generalisation to the broader ageing population should be made with caution. Competing risk of death was not modelled, as Fine-Gray approaches cannot incorporate survey weights, and our primary aim was to estimate direct associations, for which cause-specific hazard models are more appropriate,^45^ rather than individual risk prediction. Residual confounding such as frailty or genetic susceptibility cannot be ruled out.

In conclusion, this study identifies domain-specific, bidirectional links between infection and cognition. Severe infections were most consistently associated with poorer executive function, with LRTI showing an additional association with memory. Conversely, multidomain cognitive deficits predicted higher risk of subsequent all-cause and bacterial infections, whereas only executive function predicted future viral infections, particularly COVID-19. These findings highlight the importance of domain-specific cognitive assessment beyond global measures when examining infection-related risk, and suggest that proactive infection-prevention strategies tailored to specific cognitive vulnerabilities may help reduce both infection risk and its downstream cognitive consequences.

## Data Availability

All data produced are available online at

https://datacatalogue.ukdataservice.ac.uk/studies/study/8502

## Consent statement

The ELSA-HCAP study was conducted in accordance with the Declaration of Helsinki, and ethical approval and experimental protocols were granted by the South Central—Berkshire Research Ethics Committee (REC Ref: 17/SC/0254). All participants gave their informed consent to take part in the study.

