## Supplementary materials for "Severe infections, domain-specific cognitive vulnerability, and future infection risk in older adults"

**Figure S1**. Flow of participants in the analytic sample

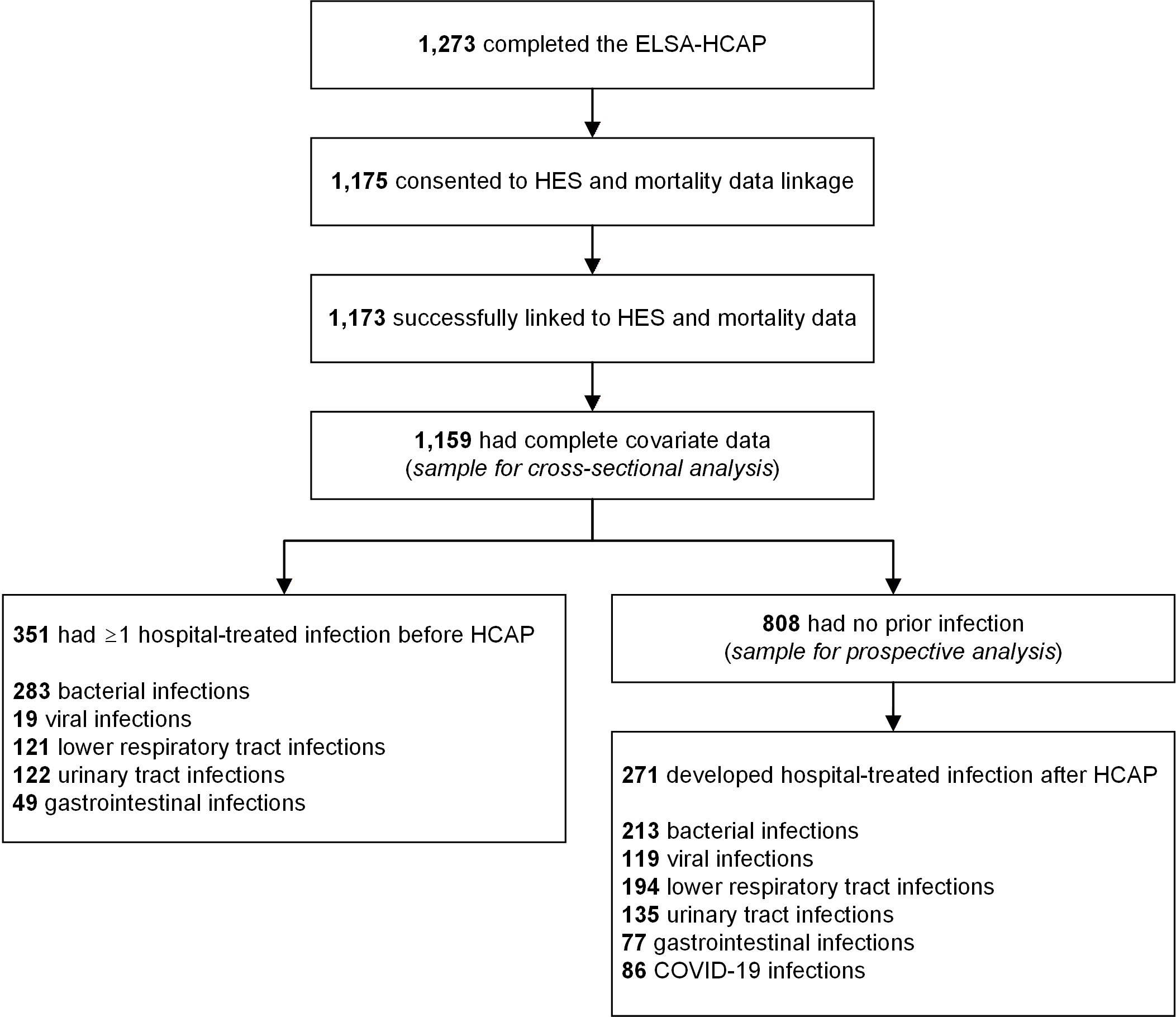

ELSA, English Longitudinal Study of Ageing; HCAP, Harmonised Cognitive Assessment Protocol; HES, Hospital Episode Statistics

**Table S1**. Cognitive test items, domains, descriptions, and missingness in ELSA-HCAP

| Test | Item | Domain | Brief description | Missing proportion |
| --- | --- | --- | --- | --- |
| MMSE | Object naming | Language | Name common objects (e.g., watch, pencil) | 1/1273 (0.1%) |
| MMSE | Command following | Language | Read and follow a written command (“close eyes”) | 14/1273 (1.1%) |
| MMSE | Phrase repetition | Language | Repeat a spoken phrase | 8/1273 (0.6%) |
| MMSE | Sentence writing | Language | Write a complete sentence | 44/1273 (3.5%) |
| MMSE | Serial 7 subtraction | Executive Functioning | Subtract 7s serially from 100 | 97/1273 (7.6%) |
| MMSE | Recall | Memory | Recall 3 words immediately | 5/1273 (0.4%) |
| MMSE | Recall | Memory | Recall 3 words after delay | 8/1273 (0.6%) |
| HRS-TICS | Vocabulary & factual knowledge | Language | Identify words (e.g., scissors, cactus) and name UK Prime Minister | 8/1273 (0.6%) |
| CERAD Word List | Immediate recall | Memory | Recall 10 words across three trials | 10/1273 (0.8%) |
| CERAD Word List | Delayed recall | Memory | Recall the 10 words after a delay | 9/1273 (0.7%) |
| CERAD Word List | Recognition | Memory | Recognize 10 original words among distractors | 12/1273 (0.9%) |
| CERAD Constructional Praxis | Immediate drawing | Visuospatial | Copy geometric figures (circle, cube, diamond, rectangles) | 68/1273 (5.3%) |
| CERAD Constructional Praxis | Delayed recall drawing | Memory | Redraw shapes from memory | 7/1273 (0.5%) |
| Animal Naming | Animal retrieval fluency | Language | Name as many animals as possible in 60s | 3/1273 (0.2%) |
| WMS-IV | Brave Man story – immediate | Memory | Recall story details immediately | 9/1273 (0.7%) |
| WMS-IV | Brave Man story – delayed | Memory | Recall story details after delay | 56/1273 (4.4%) |
| WMS-IV | Robbery story – immediate | Memory | Recall story details immediately | 20/1273 (1.6%) |
| WMS-IV | Robbery story – delayed | Memory | Recall story details after delay | 63/1273 (4.9%) |
| WMS-IV | Robbery story – recognition | Memory | Recognize true/false statements about story | 51/1273 (4.0%) |
| Ps & Ws Letter Cancellation | Letter cancellation | Executive Functioning | Cross out Ps and Ws on a letter grid in 60s | 87/1273 (6.8%) |
| Backward Counting | Backward counting | Executive Functioning | Count backwards from 100 for 30s | 20/1273 (1.6%) |
| SDMT | Symbol-digit substitution | Executive Functioning | Pair symbols with digits under time pressure | 77/1273 (6.0%) |
| Number Series | Numeric reasoning | Executive Functioning | Solve incomplete numeric sequences | 120/1273 (9.4%) |
| Raven's progressive matrices | Pattern reasoning | Executive Functioning | Identify missing shapes in abstract visual patterns | 15/1273 (1.2%) |
| Trail Making Test | Part A | Executive Functioning | Connect numbered circles in order (speed test) | 64/1273 (5.0%) |
| Trail Making Test | Part B | Executive Functioning | Alternate connecting numbers and letters | 235/1273 (18.5%) |
| CSI-D | Naming Items | Language | Object naming, usage, directions, pointing | 2/1273 (0.2%) |
| Other tasks | Paper-folding (3-step) | Language | Follow a three-step paper-folding instruction | 5/1273 (0.4%) |
| Other tasks | Interlocking shapes | Visuospatial | Draw overlapping/interlocking figures | 21/1273 (1.6%) |

MMSE, Mini-Mental State Examination; HRS-TICS, HRS Telephone Interview for Cognitive Status; CERAD, Consortium to Establish a Registry for Alzheimer's Disease; WMS-IV, Wechsler Memory Scale IV; SDMT, Symbol Digit Modalities Test; CSI-D, Community Screening Interview for Dementia. Proportions are unweighted.

**Table S2**. ICD-10 codes used to define covariates.

| Disease | ICD-10 code |
| --- | --- |
| Diabetes | E10, E11, E13 |
| Hypertension | I10, I11, I12, I13, I15 |
| Cardiovascular disease | I20, I21, I22, I23, I24, I25, I50, I60, I61, I63, I64 |
| Chronic lung disease | J41, J42, J43, J44 |
| Psychiatric condition | F20–F48 |

ICD-10, International Classification of Diseases, Tenth Revision. List of diseases included as covariates, with the corresponding ICD-10 codes used to identify diagnoses from Hospital Episode Statistics (HES) records.

**Table S3**. Definitions, categorizations, and data sources for covariates used in the analyses

| Variable | Data source | Definition / categorization |
| --- | --- | --- |
| Age | HCAP | Interview year - birth year |
| Sex | HCAP |  |
| Living status | ELSA | Number of people in household: 1) Alone; 2) >1 Co-residing |
| Education | ELSA (IFS derived) | Educational qualification (information merged from current and previous waves): 1) <O level; 2) O level; 3) ≥A level |
| Wealth | ELSA (financial derived) | Benefit unit total net (non-pension) wealth (in quintiles). “Benefit unit” = single adult or couple living as married and dependent children |
| Index of Multiple Deprivation | ELSA | Small-area socioeconomic measure (~1,500 residents) combining income, employment, education, health, crime, barriers to housing and services, and living environment. In quintiles |
| Loneliness | ELSA | Assessed using three questions on how often participants felt a lack of companionship, left out, or isolated from others. Responses (“hardly ever or never,” “some of the time,” “often”) were summed to give a score from 3 to 9 (≥6 indicating loneliness) |
| Smoking status | ELSA (IFS derived) | Cigarette smoking status: 1) Never; 2) Past/current (occasional, regular, unknown frequency ex-smoker, or current smoker) |
| Alcohol use | ELSA | Alcohol consumption in past 12 months: 1) ≥5 days/week (Almost every day; five or six days a week); 2) <5 days/week (Three or four days a week; once or twice a week; once or twice a month; once every couple of months; once or twice a year; not at all) |
| Diabetes | ELSA, HCAP, HES | Self-reported diabetes/high blood sugar or HES diagnosis (ICD codes in Table S2) |
| Hypertension | ELSA, HCAP, HES | Self-reported high blood pressure/hypertension or HES diagnosis (ICD codes in Table S2) |
| Cardiovascular disease | ELSA, HCAP, HES | Self-reported angina, heart attack, heart failure, or stroke; or HES diagnosis (ICD codes in Table S2) |
| Chronic lung disease | ELSA, HCAP, HES | Self-reported chronic lung disease (e.g., bronchitis, emphysema, chronic obstructive pulmonary disease) or HES diagnosis (ICD codes in Table S2) |
| Psychiatric condition | ELSA, HCAP, HES | Self-reported emotional, nervous or psychiatric problems or HES diagnosis (ICD codes in Table S2) |

ELSA, English Longitudinal Study of Ageing; HCAP, Harmonised Cognitive Assessment Protocol; IFS, Institute for Fiscal Studies; HES, Hospital Episode Statistics; ICD, International Classification of Diseases.

**Table S4**. Missing data for covariates

| Covariates | Missing (n) | Missing (%) ^a^ |
| --- | --- | --- |
| Age | 0 | 0 |
| Sex | 0 | 0 |
| Living status | 0 | 0 |
| Loneliness | 9 | 0.8 |
| Education | 1 | 0.1 |
| Wealth | 4 | 0.3 |
| Index of Multiple Deprivation | 0 | 0 |
| Smoking status | 0 | 0 |
| Alcohol use | 1 | 0.1 |

^a^ Denominator = 1,180; Proportions are unweighted.

**Table S5**. Associations between any pre-HCAP hospital-treated infection and domain-specific cognitive factor scores

| Domain | Model 1 | Model 2 | Model 3 |
| --- | --- | --- | --- |
| General cognition | -0.22 (-0.32 to -0.12) | -0.17 (-0.26 to -0.08) | -0.11 (-0.21 to -0.02) |
| Memory | -0.11 (-0.22 to -0.00) | -0.08 (-0.18 to 0.03) | -0.05 (-0.16 to 0.06) |
| Language | -0.13 (-0.24 to -0.02) | -0.10 (-0.21 to 0.01) | -0.05 (-0.16 to 0.06) |
| Visuospatial ability | -0.14 (-0.26 to -0.02) | -0.09 (-0.20 to 0.02) | -0.03 (-0.15 to 0.08) |
| Executive function | -0.28 (-0.38 to -0.18) | -0.24 (-0.33 to -0.14) | -0.19 (-0.28 to -0.09) |

Values are β coefficients (95% confidence intervals) from multivariable linear regression models estimating differences in standardized cognitive factor scores associated with any hospital-treated infection before HCAP. Model 1 adjusted for age and sex. Model 2 additionally adjusted for socioeconomic and lifestyle factors. Model 3 further adjusted for health-related variables. Negative values indicate lower cognitive performance associated with prior infection. All models were weighted to account for differential sampling and participation probabilities at HCAP.

**Figure S2**. Schoenfeld residuals for general cognition in the Cox proportional hazards model

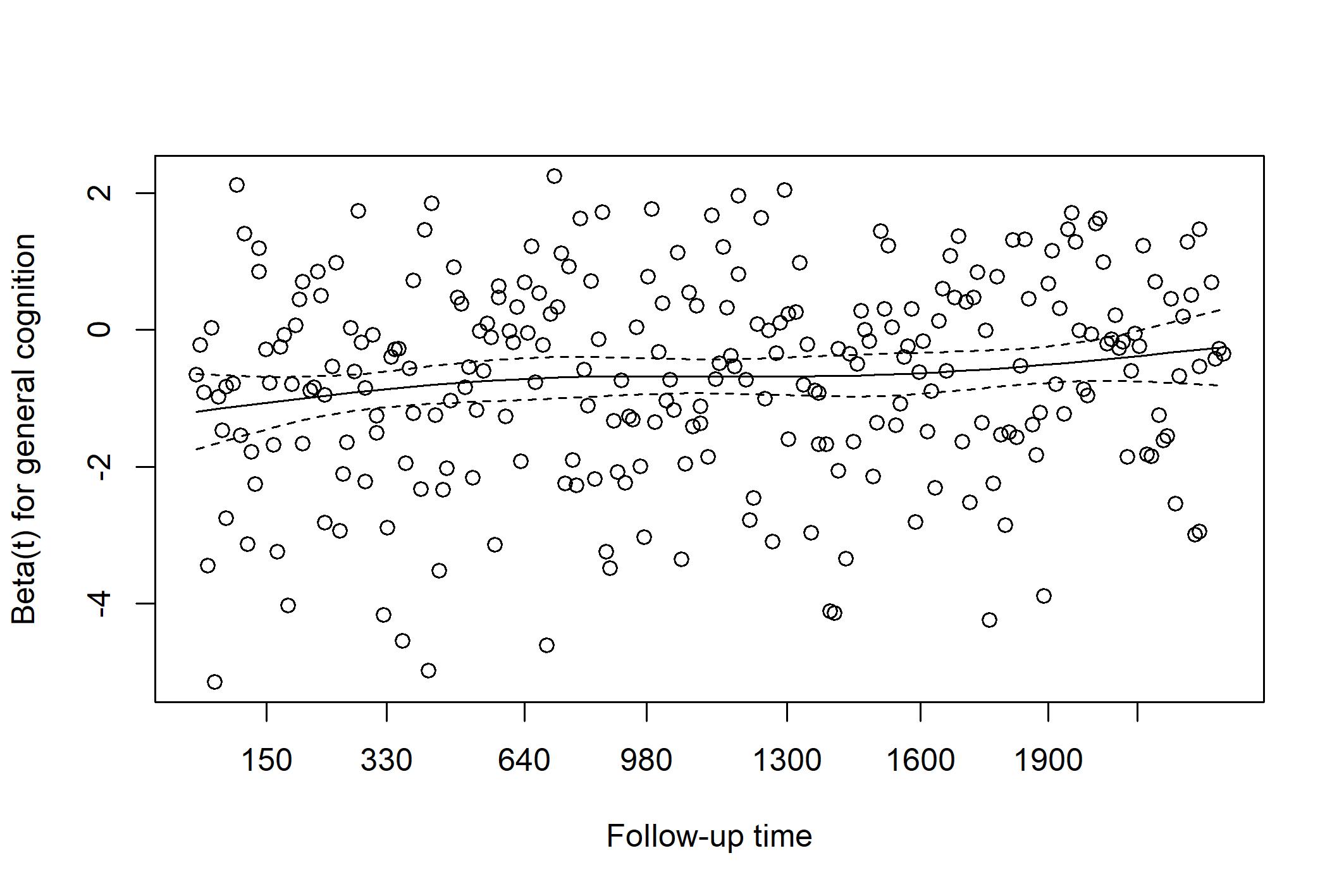

Schoenfeld residuals from the Cox model assessing the association between standardized general cognitive factor scores and subsequent hospital-treated infection. The smoothed estimate (solid line) and its confidence bands (dashed lines) show no systematic deviation from a horizontal trend over follow-up time (days), indicating no evidence of violation of the proportional hazards assumption for general cognition.

**Table S6**. Associations between cognitive factor scores at HCAP and risk of subsequent incident hospital-treated infection with sequential covariate adjustment

| Domain | Model 1 | Model 2 | Model 3 |
| --- | --- | --- | --- |
| General cognition | 0.63 (0.51 to 0.76) | 0.63 (0.52 to 0.77) | 0.64 (0.53 to 0.78) |
| Memory | 0.70 (0.58 to 0.84) | 0.72 (0.60 to 0.86) | 0.71 (0.60 to 0.86) |
| Language | 0.72 (0.61 to 0.86) | 0.75 (0.63 to 0.89) | 0.77 (0.65 to 0.93) |
| Visuospatial ability | 0.82 (0.70 to 0.97) | 0.88 (0.75 to 1.05) | 0.89 (0.76 to 1.05) |
| Executive function | 0.65 (0.54 to 0.79) | 0.66 (0.55 to 0.80) | 0.67 (0.56 to 0.82) |

Values are hazard ratios (HRs) with 95% confidence intervals from multivariable Cox proportional hazards models estimating associations between standardized cognitive factor scores measured at HCAP and risk of incident hospital-treated infection.

Model 1 adjusted for age and sex; Model 2 additionally adjusted for socioeconomic and lifestyle factors; Model 3 further adjusted for health-related variables. HRs are expressed per 1 SD higher cognitive score, with values <1 indicating lower infection risk. All models were weighted to account for differential sampling and participation probabilities at HCAP.

**Figure S3**. Association between HCAP general cognitive function and risk of subsequent hospital-treated infection by subgroup

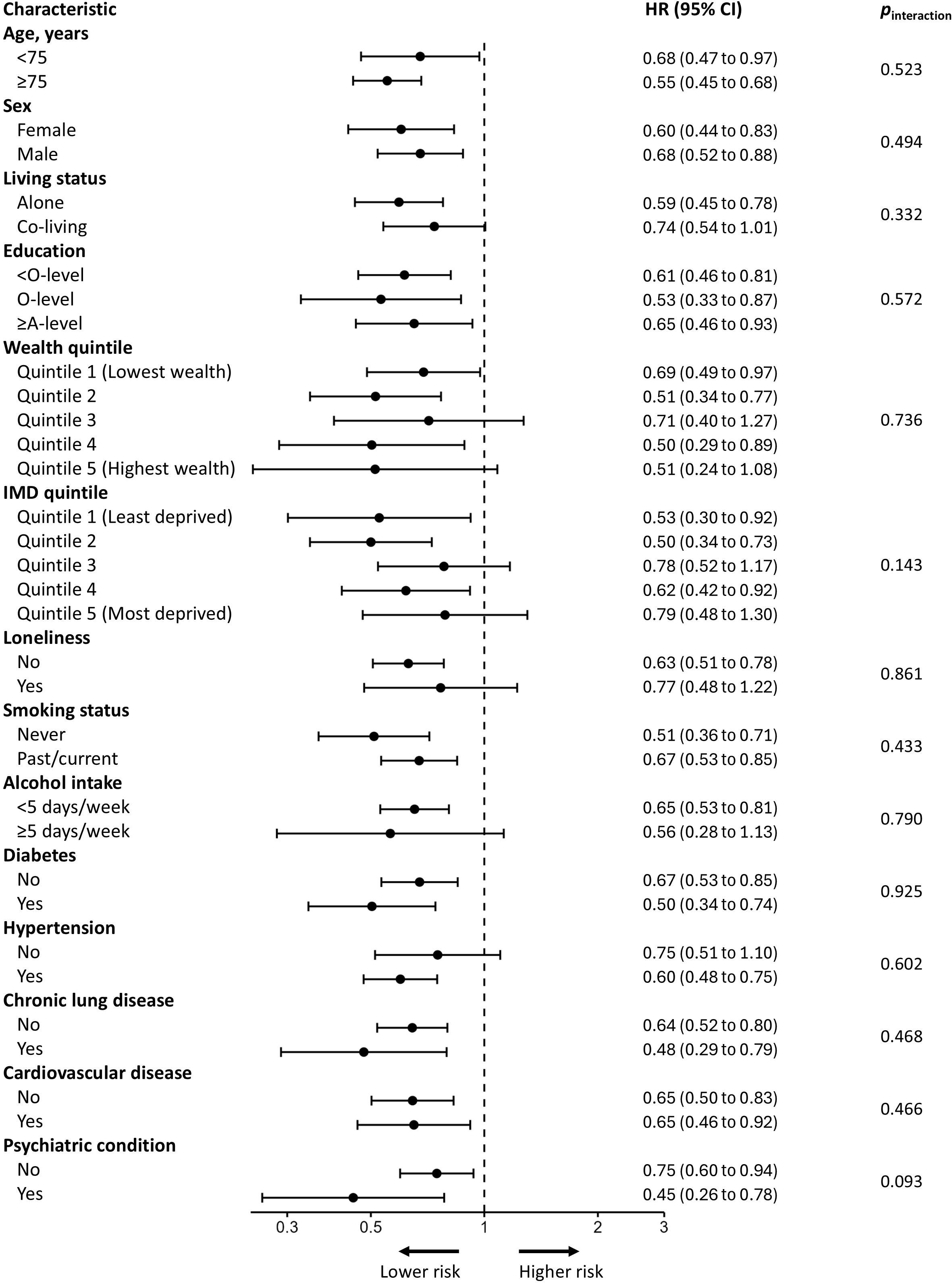

CI, confidence intervals, HR, hazard ratio; IMD, Index of Multiple Deprivation. Associations of standardized general cognitive factor scores measured in HCAP with risk of subsequent hospital-treated infection, stratified by demographic, socioeconomic, lifestyle, and health characteristics. Models were adjusted for all listed covariates and weighted to account for differential sampling and participation probabilities at HCAP. p_interaction_ values were obtained using likelihood ratio tests.
